# Relationships between alcohol use and dementia: protocol for an observational study in the UK Clinical Practice Research Datalink

**DOI:** 10.1101/2025.10.09.25337654

**Authors:** Nasri Fatih, Krishnan Bhaskaran, Aden Chun Hei Kwok, Klaus P Ebmeier, Thomas E Nichols, Joel Gelernter, Maria D. Christodoulou, Anya Topiwala

## Abstract

**Introduction:** Alcohol consumption is an increasingly recognised modifiable risk factor for dementia, yet whether it has differential impacts on dementia subtypes and its role in disease progression remains unclear. This study aims to: 1) quantify the association between alcohol intake and incidence of dementia subtypes, and 2) examine whether individuals who drink heavily and develop dementia referred to herein as ‘alcohol-related’ - have poorer post-diagnosis outcomes compared to other dementia cases. Clarifying these relationships will determine whether alcohol selectively increases risk for specific dementia phenotypes or broadly heightens neurodegenerative vulnerability, with implications for prevention, clinical counselling, and therapeutic targeting.

**Methods and analysis:** This population-based cohort study will use linked UK electronic health records from Clinical Practice Research Datalink (CPRD), Hospital Episode Statistics (HES), and Office for National Statistics (ONS). Alcohol exposure will be defined through self-reported recorded weekly alcohol units and diagnostic codes for harmful or dependent alcohol use. Primary outcomes including incident all-cause and subtype-specific dementia (e.g., Alzheimer’s, vascular, Lewy body, Parkinson’s, frontotemporal) as well as secondary outcomes (i.e., mortality, care-home entry, and neuropsychiatric symptoms). Key covariates encompassing sociodemographic factors, smoking, and relevant comorbidities will be adjusted for. Multivariable Cox proportional hazards and Fine–Gray competing risk models will estimate associations with dementia incidence. Post-diagnosis prognosis will be compared for dementia in individuals with a history of heavy alcohol use (‘alcohol-related’) and dementia in individuals with minimal alcohol exposure (‘non-alcohol-related’) cases using survival and logistic regression models. Multiple testing correction will be applied across dementia subtype comparisons. Alcohol exposure will be modelled continuously and nonlinearly using restricted cubic splines and categorically using binary indicators of harmful/dependent use. Missing covariate data will be assessed and addressed using appropriate methods, including multiple imputation and complete-case analysis. Data extraction and analysis are scheduled from 10/2025 to 10/2026.

**Ethics and dissemination:** Use of deidentified routine data will proceed under existing Research Ethics Committee and data governance approvals. Findings will be disseminated via open-access peer reviewed journals, academic conferences, and summaries targeted at patient, public and policy audiences. The results of this study will be reported according to the STROBE and RECORD guidelines.

**Article Summary:** *Strengths and Limitations of this study:*

- This study uses linked CPRD, HES, and ONS data, providing a large, nationally representative UK sample with improved capture across the full alcohol exposure range, particularly of individuals with hazardous or dependent alcohol use, who are often underrepresented in biobank studies.
- The use of prospectively collected, long-term data enables temporal relationships between alcohol exposure and dementia onset to be examined, helping to minimise reverse causation and allowing for trajectory analysis over time.
- Dementia case ascertainment is strengthened by combining data from primary care, hospital, and death records.
- Alcohol intake data are self-reported and subject to measurement error. Missing data on alcohol intake is a significant limitation and may impact statistical power and generalisability. Random measurement error is likely to bias findings conservatively towards the null.
- As with all observational studies, residual confounding remains a risk despite planned adjustment for key covariates. While electronic health records are increasingly validated for dementia diagnoses, subtyping may be less reliable. Findings will be triangulated with analyses in other cohorts using alternative dementia phenotyping methods.

## Introduction

Alcohol-related harm is expected to rise as populations age and alcohol consumption remains widespread (1,2). While heavy, prolonged drinking is an established risk factor for cognitive decline and alcohol-related dementia including Korsakoff’s syndrome, (3–6) accumulating evidence indicates that even moderate intake may adversely affect brain structure and function (6–8). Given the difficulties finding disease-modifying treatments for dementia, prevention is an important strategy. Effective prevention strategies necessitate a deeper understanding of the mechanistic relationship between alcohol and dementia (9). It remains unclear whether alcohol selectively increases risk of certain dementia types or exerts a more general neurodegenerative effect. Alcohol may act through multiple, partially overlapping pathways—vascular injury, neuroinflammation, thiamine deficiency, metabolic dysregulation, direct neurotoxicity (10). Disentangling subtype-specific associations may offer insight into these underlying biological pathways. Moreover, alcohol’s effects on dementia may vary by factors such as sex, yet such interactions have not been well-characterised. Drinking history may also influence prognosis after dementia diagnosis, including survival, institutionalisation, and neuropsychiatric symptom burden. Clarifying these relationships may inform prevention strategies, enhance clinical counselling, and identify opportunities for therapeutic repurposing.

### Objectives

#### Primary

1. To estimate dose-response relationships between alcohol use (quantity and harmful/dependent use) and incident dementia overall and by subtype (Alzheimer’s disease, vascular dementia, Lewy body dementia, Parkinson’s disease dementia, frontotemporal dementia).
2. To assess heterogeneity of alcohol-dementia associations by demographic and clinical strata (e.g., sex, deprivation quintiles, smoking status).

### Secondary

1. To examine whether prognosis following dementia diagnosis (time to death/ institutionalisation, neuropsychiatric symptom prevalence) differs between alcohol-related and non–alcohol-related dementia.
2. To examine associations between alcohol and early-onset vs late-onset dementia

#### Hypotheses

- H1: Higher alcohol intake and clinically harmful/dependent use will be associated with increased incidence of both common and rarer dementia subtypes..
- H2: Alcohol-dementia associations will differ by demographic and clinical strata with evidence suggestive of vulnerability in women and in those with lower socioeconomic status.
- H3: Individuals with alcohol-related dementia experience shorter times to death and institutionalisation, and higher odds of neuropsychiatric symptoms, than those with non-alcohol-related dementia.
- H4: Higher alcohol consumption is likely to be associated with early- and late-onset dementia.

## Methods and analysis

### Patient and public involvement

Patients and the public were not involved in the design, conduct, reporting, or dissemination plans of this research.

### Study design and setting

This is a retrospective, population-based cohort study using routinely collected electronic health records from general practices in the United Kingdom contributing to the Clinical Practice Research Datalink (CPRD). CPRD is a large, nationally representative database containing anonymised primary care records, covering approximately 20-25% of the UK population (11,12). The dataset is broadly representative in terms of age, sex, and ethnicity to the wider UK population (13,14). For this study, CPRD data will be linked at the individual level to:

1. **Hospital Episode Statistics (HES)**: providing information on inpatient admissions, diagnoses, and procedures and ethnicity for NHS patients in England.
2. **Office for National Statistics (ONS) mortality data:** providing dates and certified causes of death. Index of Multiple Deprivation (IMD) for person-level proxy of socioeconomic status.

Linkage is available for patients registered with practices that have consented to data linkage. The study setting therefore encompasses a real-world UK population receiving care across both primary and secondary care sectors, enabling comprehensive longitudinal assessment of clinical exposures, comorbidities, and outcomes.

### Exposures

As outlined in Table 1, alcohol use will be ascertained in two ways: (1) recorded weekly alcohol units (recorded both continuously and categorically); and (2) diagnostic codes indicating harmful or dependent use codes, derived from primary care (Read codes) and secondary care (ICD-10 codes in HES). Code lists ascertained from existing libraries (e.g. HDR phenotype and London School of Hygiene and Tropical Medicine website) and the CPRD code browser were reviewed by a psychiatrist.

**Table 1.** Definition of the Study Population with inclusion and exclusion criteria.

| Aspect | Definition |
| --- | --- |
| **Target Population** | Adults in CPRD GOLD and Aurum with linkage to HES and ONS where available. |
| **Recruitment / Follow-up Period** | **Start /Index date:** earliest date of alcohol intake recording.<br>**End:** from index date + 1-year lag until earliest of dementia diagnosis, death, transfer out of practice, or end of data collection. |
| **Inclusion Criteria** | Permanently registered with a CPRD practice<br>Age $\geq 40$ at index date.<br>Acceptable patient flag in CPRD.<br>Eligible for linkage to HES/ONS (if using linked outcomes). |
| **Exposure Definition** | **Primary objectives:** alcohol units per week (continuous) or categorical (e.g. light drinker <7 units).<br>**Secondary objectives:** participants are grouped into alcohol-related dementia and non-alcohol related dementia on various criteria.<br>- **Alcohol-related dementia\*:** evidence of an alcohol-dementia diagnosis as per read/ICD codes OR dementia code plus 50 units of alcohol per week OR a read/ICD code of alcohol use disorder.<br>- **Non-alcohol-related dementia:** Dementia diagnosis but no evidence of an alcohol-dementia diagnosis in read/ICD codes < 7 units of alcohol per week and no evidence of a read/ICD code of alcohol use disorder. |
| **Outcomes** | **Primary objectives:** Incident dementia (all-cause).<br>**Secondary objectives:** Alcohol-related dementia, other dementia subtypes, time to death/institutionalisation, neuropsychiatric symptoms (in dementia cases). |
| **Exclusions** | Patients will be excluded if they do not have valid, continuous registration and plausible demographics (as per CPRD acceptable criteria) or have a year of birth before death or registration and institutionalisation at baseline.<br>*Prevalent exclusions:* exclude individuals with any dementia/cognitive-impairment code before the index date.<br>*Latency (immortal time) handling:* apply a 1-year lag after the index date; no person-time or events within this window contribute to the analysis. |

\*We use the term 'alcohol-related dementia' to describe dementia observed in individuals with a history of heavy alcohol consumption, while acknowledging that multiple other etiological factors also contribute to dementia risk.

To assess the validity of alcohol exposure data, we will compare the distribution of recorded alcohol intake - stratified by age and sex - to external, nationally representative datasets such as the Health Survey England. We will also conduct negative and positive control analyses (as described below) to evaluate the potential for nondifferential misclassification bias due to missing or misreported alcohol use.

Alcohol-related dementia will be defined by the presence of any dementia code (including Wernicke-Korsakoff’s syndrome), or the presence of any harmful/dependent alcohol code, or recorded units per week greater than 50 units (defined empirically) or 35 units for women as per the Oslin clinical guidelines (15).

Non-alcohol-related dementia will be defined by the presence of any dementia code and the absence of a harmful or dependent alcohol code, and recorded alcohol units less than 7 units per week.

### Outcomes

Primary and secondary outcomes will be derived using available read and ICD codes in primary care, secondary care and death records. Primary outcomes include: incident all-cause dementia and subtypes (Alzheimer’s disease, vascular, Lewy body, Parkinson’s disease dementia, frontotemporal/Pick’s). When dementia is identified only on the death certificate, we will classify it as incident dementia with the date of death used as the event date. Secondary outcomes include: date of death (from ONS linked data), residing in a care home as recorded either in primary care (read code of event or observation date) or HES (admitted/discharged to a care home), presence of neuropsychiatric symptoms (agitation, aggression, depression, anxiety, apathy or psychosis read codes and/or psychotropic medication prescription).

### Covariates

Relevant confounds have been identified from the literature (16–19). A directed acyclic graph (DAG) underpins our modelling approach, identifying age, socioeconomic deprivation, smoking, ethnicity and BMI as key confounds with available data in this dataset. Age at baseline will be calculated by subtracting date at alcohol measurement from year of birth. ONS-derived ethnicity is presented as self-reported in categories (Asian, Black, mixed/multiple, white, other and unknown.) Smoking status, recorded as a categorical variable (current smoker, ex-smoker and never smoked) and BMI (identified through read codes or calculated from height and weight) will be ascertained as temporally proximal to the alcohol measure as possible using previously published codes (20). Socioeconomic status will be measured using the 2019 Index of Multiple Deprivation (IMD), an area-level indicator based on seven domains (income, employment, education, health, crime, housing, environment). Patient postcodes were linked to Lower Layer Super Output Areas (LSOAs), and each assigned an IMD decile (1 = least deprived, 10 = most deprived) to capture relative deprivation. Other important confounders of the relationship examined such as diet, education and physical activity were not measured in the dataset and thus unable to be considered. Liver disease, cardiovascular or cerebrovascular events and traumatic brain injury (captured through clinical codes) will not be considered as confounders but, in later analyses, will be examined as potential mediators of the alcohol-dementia associations.

### Sample size calculation

With an anticipated cohort size of 1,000,000 participants with alcohol data, and assuming a 5% prevalence of heavy drinking, we estimate 90% power to detect a dementia hazard ratio of 1.02 at α = 0.05 (prior to correction for multiple testing). In addition, a case–control design including 50,000 alcohol-related dementia cases and 50,000 age-matched dementia controls provides 80% power to detect a hazard ratio of 1.02.

### Data analyses plan/statistical methods

For our first objective, alcohol exposure will be examined both as a continuous measure (units per week, standardised from daily/monthly reports) and as a categorical measure (light, moderate, heavy, ex- and non-drinkers). Restricted cubic splines (5 knots) will be fitted to the continuous measure to assess non-linear associations. Associations with dementia incidence will be estimated using Cox proportional hazards models adjusting for covariates. Time at risk will be calculated as the interval from the first valid alcohol record until dementia diagnosis or censoring (death or end of follow-up). Proportional hazards assumptions will be formally tested with Schoenfeld residuals, with violations addressed by stratification or time-varying terms. Given that our primary causal question concerns dementia onset, deaths without dementia will be censored; however, Fine–Gray subdistribution hazards models will additionally be used to provide cumulative incidence estimates accounting for competing risk of death (21).

For our secondary objective, we will examine whether there are clinical differences between the alcohol-related and non-alcohol related dementia groups. Logistic regression models will be used to assess whether date of death (from ONS linked data), institutionalisation as recorded either in primary care (read code of event or observation date) or HES (admitted/discharged to a care home), neuropsychiatric symptoms (agitation/aggression, psychosis read codes and/or anti-psychotic prescription differ between the two groups.

Quantitative bias analysis will be used to evaluate potential bias due to unmeasured confounding or misclassification. Patterns of missing data will be examined with respect to observed variables, and appropriate methods (i.e., multiple imputation or delta-adjusted complete-case analysis) used to handle missingness. Multiple testing correction will be applied using false discovery rate when comparing dementia subtypes.

Several sensitivity and robustness analyses will be considered. These include use of negative controls: (i) Outcome: e.g., dementia; (ii) Exposure: recorded eczema diagnosis or seasonable allergy meds (plausibly unrelated to dementia pathways after adjustment) to probe residual systematic bias and the existence of a spurious association. Established alcohol-related diseases such as liver cirrhosis (22) will be used as positive controls to test whether associations follow expected patterns thereby providing confidence that our analytic approach can recover known causal relationships (e.g., between heavy alcohol use and cirrhosis) and strengthen confidence in the validity of any observed associations with dementia.

Missingness patterns in alcohol data within CPRD will be systematically examined with respect to recorded sociodemographic factors, including age, sex, ethnicity, deprivation level and smoking status. Missingness amongst covariate data will be thoroughly examined and approaches such as multiple imputation by chained equations (MICE) under missing-at-random (MAR) assumptions and complete-case with delta-adjusted sensitivity analyses to assess missing-not-at-random (MNAR) impact will be considered. Family-wise error will be controlled using Holm–Bonferroni for primary subtype comparisons. False discovery rate (Benjamini–Hochberg) will be applied to secondary endpoints.

### Future analyses

Future analyses will model longitudinal trajectories of alcohol consumption (e.g., units per week) to examine changes over time in relation to dementia diagnosis, including the potential for reverse causation.

We also aim to investigate the mechanisms that underlie alcohol-dementia associations by examining the role of potential mediators such as liver disease, cardiovascular or cerebrovascular events and traumatic brain injury (captured through clinical codes) using causal mediation and/or structural equational modelling analyses (23,24). Effect modification by factors including sex will be explored by testing interaction terms with alcohol. Marginal effects will be presented to aid interpretation.

### Ethics, Governance and Patient/Public Involvement

This study is based in part on data from the Clinical Practice Research Datalink obtained under licence from the UK Medicines and Healthcare products Regulatory Agency. The data is provided by patients and collected by the NHS as part of their care and support. The interpretation and conclusions contained in this study are those of the author/s alone. Study reference number: 24_004516.

## Data Availability

This study uses anonymised patient data from the UK Clinical Practice Research Datalink (CPRD) linked with Hospital Episode Statistics (HES) and Office for National Statistics (ONS) mortality data. These data are not publicly available but can be accessed by qualified researchers via application to the Independent Scientific Advisory Committee (ISAC).

https://www.cprd.com/research-applications

## Dissemination and Impact Plan

Findings will be disseminated through: (1) open-access peer-reviewed publications (e.g., target: BMJ, Lancet Public Health), (2) conference presentations (Alzheimer’s Association International Conference, UK Dementia Congress), (3) lay summaries and infographics hosted on institutional websites, (4) policy briefs for clinicians and commissioners, and (5) openly shared code lists and analysis scripts (GitHub). Results will inform clinical guidance on alcohol counselling in dementia prevention and management and may highlight opportunities to repurpose existing dementia pharmacotherapies in alcohol-related cognitive decline.

## Author Contributions

AT and NF designed the study and drafted the initial protocol. KB and MDC were responsible for methodology development and critically revised the manuscript. AK provided methodological support on the dataset. KPB, JEN, TEN provided valuable commentary on the manuscript. All authors read and approved the final version of the protocol.

## Patient and Public Involvement statement

Patients and/or the public were not involved in the design, conduct, reporting, or dissemination plans of this research.

## Data availability statement

This study uses data from the CPRD. Data are not publicly available but may be obtained from CPRD upon approved protocol submission

